# A Canado-European external validation of the Kidney Transplant Failure Score

**DOI:** 10.1101/2024.10.31.24316511

**Authors:** Arthur Chatton, Kevin Assob Feugo, Émilie Pilote, Héloïse Cardinal, Robert W. Platt, Mireille E. Schnitzer

**Affiliations:** Faculté de Pharmacie, Université de Montréal, Montréal, QC, Canada; Département de Médecine Sociale et Préventive, École de Santé Publique de l’Université de Montréal, Université de Montréal, Montréal, QC, Canada; Département de Médecine, Faculté de Médecine, Université de Montréal, Montréal, QC, Canada; Centre de recherche du Centre hospitalier de l’Université de Montréal, Montréal, QC, Canada; Department of Epidemiology, Biostatistics and Occupational Health, McGill University, Montréal, QC, Canada; Department of Pediatrics, McGill University, Montréal, QC, Canada; Center for Clinical Epidemiology, Lady Davis Institute, Jewish General Hospital, Montréal, QC, Canada

**Keywords:** Clinical prediction model, Competing risk, Geographical validation, Graft failure, Survival analysis, Temporal validation

## Abstract

In kidney transplantation, obtaining early information about the risk of graft failure helps physicians and patients anticipate a potential return to dialysis or retransplantation. Clinical prediction models are commonly used to obtain such risk estimation, but their performance needs to be continuously evaluated in various contexts. We propose an external validation study of the *Kidney Transplant Failure Score* in a pooled sample of 3,144 patients transplanted between 2010 and 2015 in France, Belgium, Norway and Canada. This score is used at the first transplantation anniversary to predict the probability of graft failure over the following seven years. The target population was defined as adult recipients of a kidney from a neurologically deceased donor without graft failure in the first year post-transplantation. Graft failure was defined as a return to dialysis. Around 10% of patients returned to dialysis, and 12.6% died during the seven-year follow-up. The KTFS authors fitted a Cox model and then adjusted its coefficients to maximize the discrimination, yielding the KTFS final version. We evaluated the performance of the initial and final versions of the KTFS, as well as the performance of another model we developed to consider death as a competing event. All KTFS versions yielded similarly good discrimination (area under the time-dependant receiver operating curve around from 0.79 [0.76-0.82] to 0.80 [0.77-0.84]), while the discrimination-optimized one presented important miscalibration. Clinical utility, assessed through net benefit, was also the lowest for the discrimination-optimized version. Our results warn against using the current KTFS version and recommend using either the initial coefficients or the competing risk-based ones instead.

**Lay summary:** French nephrologists have used the Kidney Transplant Failure Score (KTFS) for nearly fifteen years to predict kidney graft failure eight years after the transplantation. Because predictive performance decreases over time, we first verified that the score could still predict correctly in France and also in other countries. Then, we compared the different KTFS formulas to find that the one currently used is suboptimal and should be avoided. Our findings show that the KTFS is still a reliable source of information for both kidney recipients and nephrologists when using its first version.

## Introduction

The need for kidney replacement therapies is a major public health issue, with worldwide use expected to total 5,439 million patients (3,899 to 7,640 million) in 2030.^1^ Kidney transplantation is considered the treatment of choice, improving both quality of life and life expectancy as compared to remaining on dialysis.^2,3^ Still, there is a shortage of organs available for donation. Optimizing long-term post-transplantation care is crucial to limit the need for novel transplantations. Clinical prediction models (CPMs), or prognostic scores, can provide meaningful information for nephrologists and may help involve patients in managing their disease. However, the quality of CPMs is generally poor in kidney transplantation.^4–6^ Kaboré et al.^5^ highlighted the performance and straightforward usage of the *Kidney Transplant Failure Score* (KTFS).^7^ Its use in clinical practice is the subject of a phase-IV randomized trial.^8^ The KTFS was developed in France in the early 2000s and validated in the same population.^7^ It is computed at the first anniversary of the transplantation and predicts graft survival, defined as the time to return to dialysis, up to seven years later using the following predictors: recipient biological sex, recipient age, last donor creatininemia, the number of previous transplantations, creatininemia at three and twelve months, proteinuria at twelve months and acute rejection during the first year of transplantation. The authors fitted a Cox model to obtain a first formula (hereafter *initial coefficients*) and then derived a second formula by adjusting the predictors’ coefficients to maximize the discrimination (hereafter *discrimination-optimized coefficients*). Such discrimination-optimized coefficients may threaten calibration in other populations.^9^ Furthermore, as external validation is currently lacking,^10^ its performance outside of France is unknown. Finally, the KTFS did not consider patient death as a competing risk, likely due to its relative scarcity in the original study’s source population.^5^ Not accounting for a competing risk when externally validating a CPM can overestimate the actual risk of kidney failure.^11^

The present study aims to externally validate the KTFS (with the initial, discrimination-optimized, or competing risk-based coefficients) in prospective cohorts from Europe and North America and provide up-to-date performance.

## Material and methods

### Study population

We used, for the validation, data from two sources and considered only patients transplanted between January 1, 2010 and December 31, 2015. EKiTE is a European network with seven centers from France, one Belgian, one Spanish, and the Norwegian national registry.^12^ The Spanish center was not included because data were unavailable at the moment of the study. We also included recipients from the Centre hospitalier de l’Université de Montréal (CHUM; Quebec, Canada). Therefore, patient heterogeneity was both temporal and geographical. A total of 3,144 patients (Figure 1) met the inclusion criteria defined in Foucher et al. (adult recipients of neurologically deceased donors with a functional transplant on the first anniversary of their transplantation without missing predictors).^7^ All participants gave informed consent for research at the time of transplantation, and the current study was approved by the Université de Montréal clinical ethics committee (#2023-4811). Note that the *original development sample* refers to those used in Foucher et al. to develop the KTFS.

**Figure 1:**
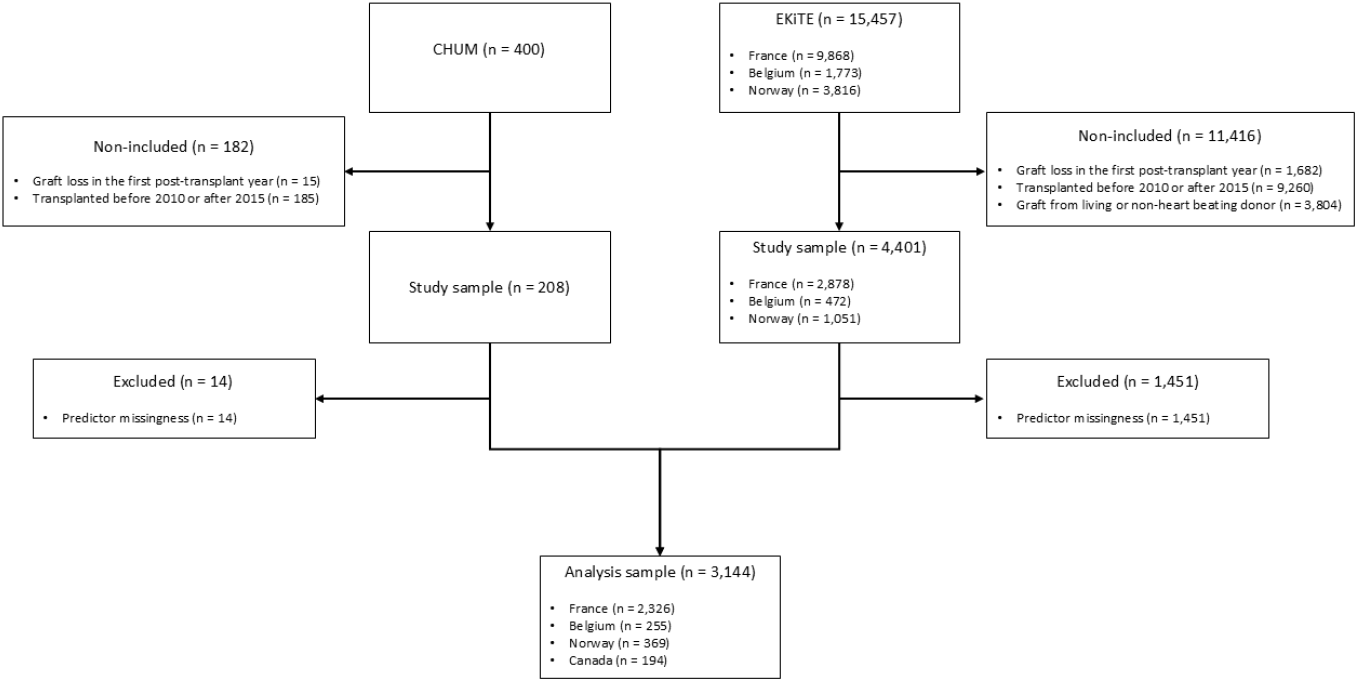
Inclusion flowchart.

### Collected Data

Donor characteristics included age, sex, and last serum creatininemia. Recipient characteristics were age, sex, body mass index, and number of previous kidney transplantations. Cold ischemia time and number of HLA incompatibilities were the transplantation-related characteristics. Post-transplantation characteristics included the occurrence of at least one acute rejection episode (including borderline changes by Banff criteria used during the study period^13,14^) during the first year posttransplantation. Finally, serum creatininemia and daily proteinuria were recorded at three, six and twelve months. Proteinuria was not collected through 24-hour urine collection on a regular basis at the CHUM during the study period, hence the urine test strip’s value was used as a proxy (value multiplied by 1.5, assuming 1.5L of urine per day on average). Delayed graft function (defined as the need for at least one dialysis session within the first-week post-transplantation) and 1-year estimated glomerular filtration rate (eGRF, estimated from the 4-variable MDRD formula^15^) were also collected. However, since French law does not authorize the storage of patient ethnicity, no recipients from French centers were considered black in the MDRD calculation.

### Outcome

The endpoint was time to graft failure, defined as the return to dialysis or retransplantation over a seven-year prediction window, with follow-up time starting one year after kidney transplantation.

### Statistical analysis

Datasets were pooled to achieve the 288 minimum number of events needed for external validation, according to Jinks et al.’s B1 formula (details on Supplementary Material A).^16^ The external validation and original development samples were compared using t-tests or Mann-Whitney tests for quantitative variables and chi-square tests for qualitative variables. Predictive performances were reported according to the Transparent Reporting of a multivariable prediction model for Individual Prognosis Or Diagnosis statement.^17^ The discrimination was evaluated by the area under the time-dependent receiver operating curve (tAUROC) at seven years.^18^ The calibration was evaluated at three different levels – mean calibration, weak calibration (calibration slope), and moderate calibration (integrated calibration index^19^) – as recommended by Van Calster et al.^9^ Overall performance was evaluated by the scaled Brier Score.^20^ Clinical utility was evaluated using a decision curve.^21^ Right-censoring was considered uninformative, and inverse probability weighting or pseudo-observation was used to take it into account.^22,23^ See McLernon et al. for more information on these metrics in survival settings.^24^ To consider the competing risk of death, the KTFS was refitted in its development database (described in Foucher et al.) using a cause-specific Cox proportional hazard model.^25^ Performance measures used were the same as above, except that the cumulative incidence functions replaced the Kaplan-Meier survival estimator.^26^ Non-parametric bootstrap (2000 iterations) was used to compute 95% confidence intervals (CIs). Finally, exploratory subgroup analyses were done at the country level using the same approaches. All analyses were conducted with R version 4.2.2.^27^

## Results

### Patient characteristics

The included patients mainly came from France (N=2,326). The sample also included 194, 255, and 369 patients in Canada, Belgium and Norway, respectively. The rate of events differed between our validation sample compared to the original development sample from Foucher et al.^7^ For instance, 10% and 12.6% of the individuals in the pooled sample presented a graft failure or a death, respectively, whereas only 8.4% and 3.6% had graft failure or death, respectively, in the development data. Survival was similar across the countries of our validation sample (Figure 2). However, the median follow-up time was longer in the validation sample than in the development data, with a range from 5.8 to 8 years for France and Canada, respectively (Table 1). This was especially visible in Canada, where more than half of the patients followed at the CHUM were administratively censored at eight years post-transplantation. The original development data’s patient characteristics differed from the pooled validation sample, even for the French subsample alone. Globally, graft overall quality was lower (e.g., older donor), while transplantation characteristics and short-term outcomes were better (e.g., shorter cold ischemia time and higher eGFR one-year post-transplantation) in the pooled validation data. This is likely explained by practice changes, such as the the use of perfusion machines.^28^

**Table 1:**
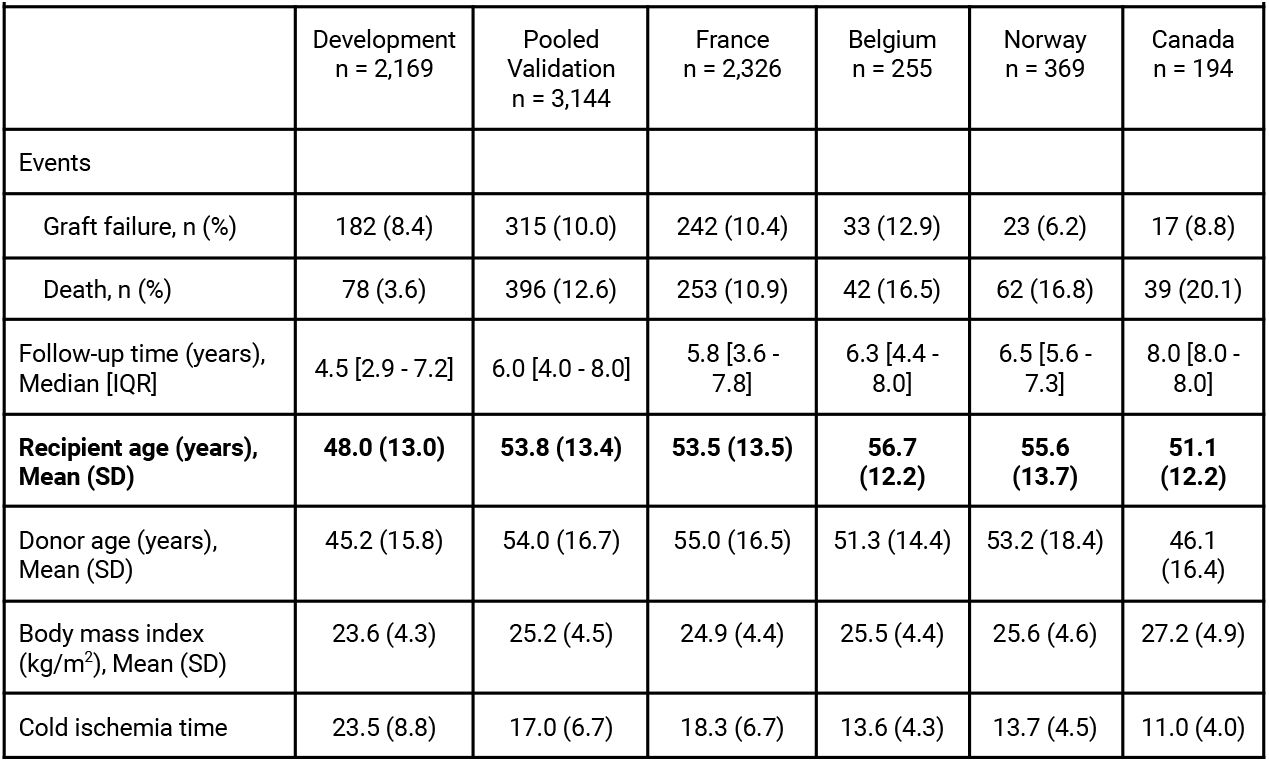
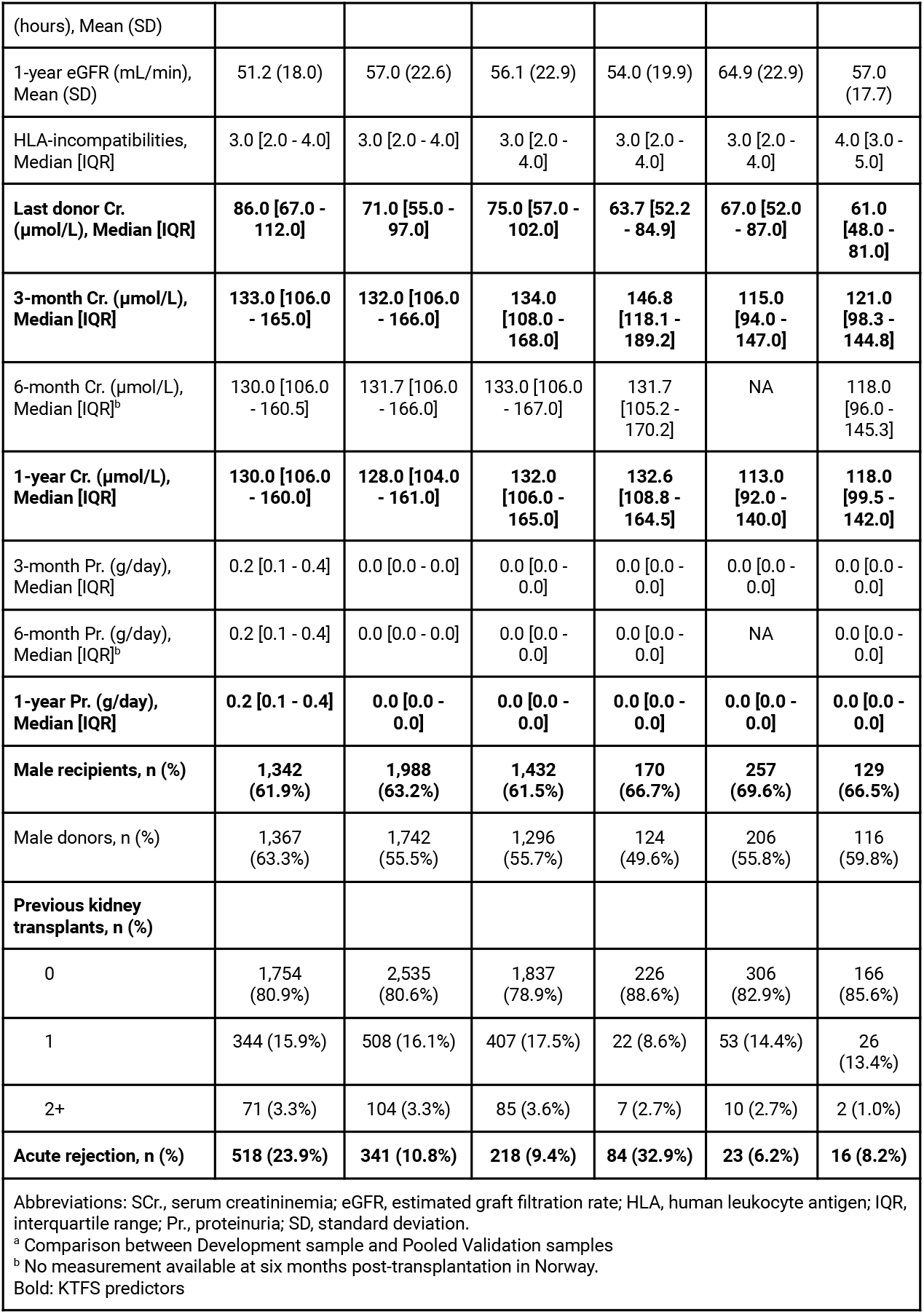
Characteristics of the patients in the initial development and validation samples (pooled or stratified by country).

**Figure 2:**
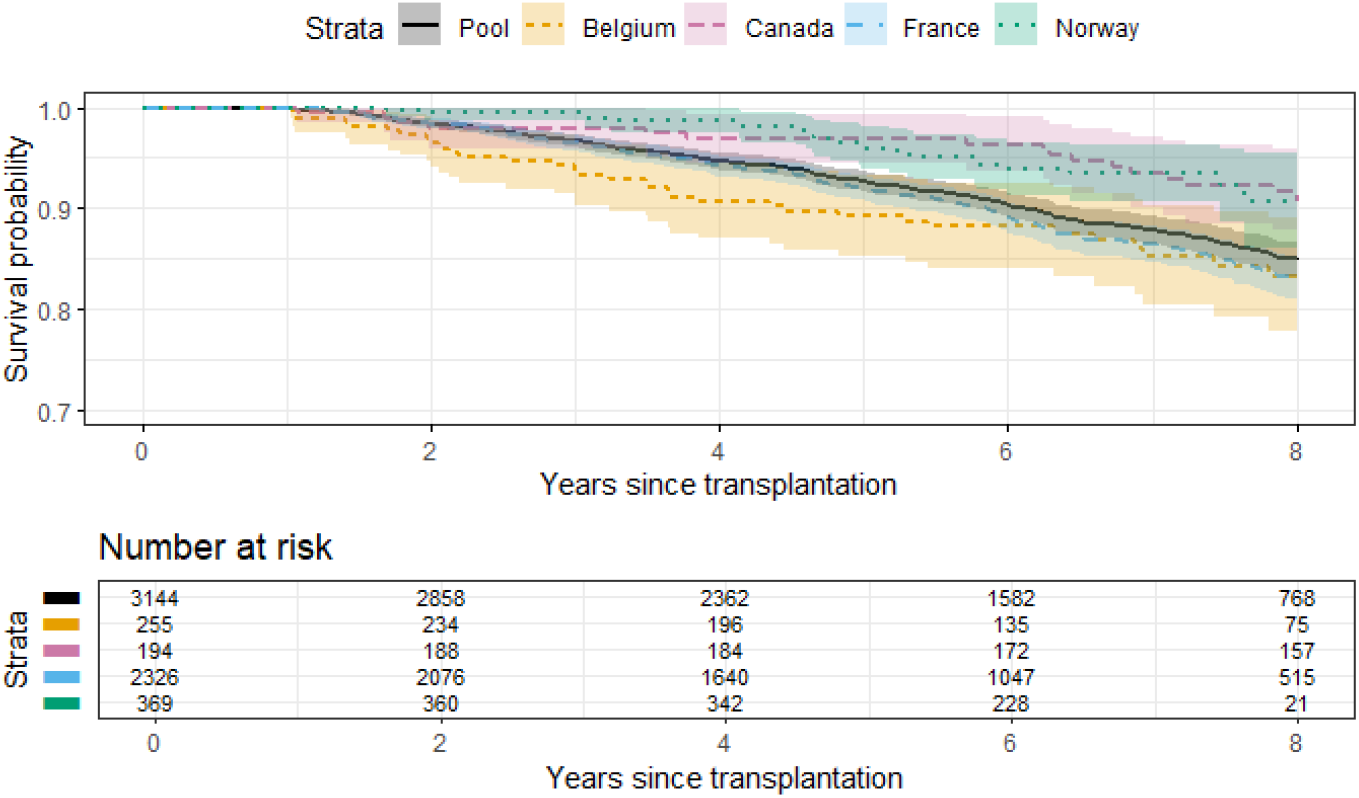
Survival curves for each country and for the pooled validation samples. Notes: The survival probability axis is trimmed before 70% to enhance between-country visibility. The sample is selected among the survivors at one year, i.e., the time of the kidney transplant failure score computation.

### Predictive capacities

The three possible KTFSs (i.e., sets of coefficients, presented in Supplementary Table S1) yielded similarly good discrimination, with a tAUROC around 0.80 (Table 2). The scaled Brier scores were also similar, meaning that approximately 20% of the prediction error of a null model was explained by the KTFS. Regarding calibration, all KTFSs showed an under-optimal weak calibration, indicating that some predictions were too extreme, and a moderate calibration close to the optimal value of zero, although it was slightly higher for the discrimination-optimized coefficients. However, the mean calibration metric showed a strong overprediction for the discrimination-optimized coefficients. See also Figure 3. There is no clear difference between the initial weights not accounting for competing risks and those considering them. Finally, Figure 4 shows increased net benefits (i.e., the “highest” line) for the KTFS over the whole range of possible thresholds. This shows that using the model to inform clinical decisions will lead to superior graft survival for any decision associated with a threshold probability of above 10% or so. Clinically speaking, a clinician thinking that missing a graft loss at seven years is three times worse than doing an unnecessary defined action (say a biopsy) will use a threshold probability of 25%.^29^ The net benefit of 0.05 at this threshold (Figure 4, panel A) means that biopsying on the basis of the KTFS is the equivalent of a strategy that identifies five graft losses at seven years per hundred recipients without conducting any unnecessary biopsy. However, the threshold chosen by the authors for being at high risk (a probability of graft failure greater than around 21.7%)^7^ showed lower net benefits on average. When using the discrimination-optimized coefficients, the net benefit of any decision associated with a KTFS threshold probability greater or equal to 60% is null.

**Table 2:**
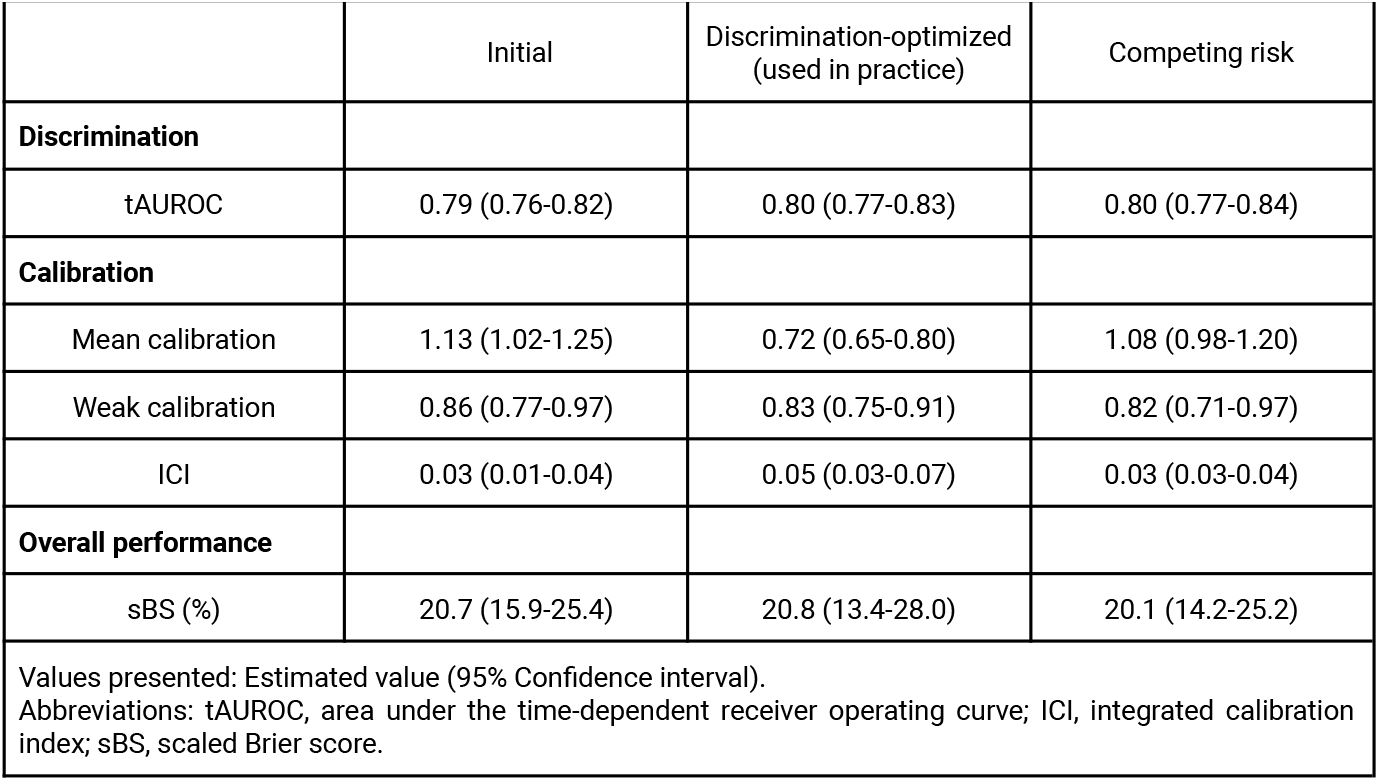
Predictive performance of the Kidney Transplant Failure Score in the pooled validation sample (N=3,144) according to the different coefficients that can be used.

|  | Initial | Discrimination-optimized<br>(used in practice) | Competing risk |
| --- | --- | --- | --- |
| **Discrimination** |  |  |  |
| tAUROC | 0.79 (0.76-0.82) | 0.80 (0.77-0.83) | 0.80 (0.77-0.84) |
| **Calibration** |  |  |  |
| Mean calibration | 1.13 (1.02-1.25) | 0.72 (0.65-0.80) | 1.08 (0.98-1.20) |
| Weak calibration | 0.86 (0.77-0.97) | 0.83 (0.75-0.91) | 0.82 (0.71-0.97) |
| ICI | 0.03 (0.01-0.04) | 0.05 (0.03-0.07) | 0.03 (0.03-0.04) |
| **Overall performance** |  |  |  |
| sBS (%) | 20.7 (15.9-25.4) | 20.8 (13.4-28.0) | 20.1 (14.2-25.2) |

Values presented: Estimated value (95% Confidence interval).
Abbreviations: tAUROC, area under the time-dependent receiver operating curve; ICI, integrated calibration index; sBS, scaled Brier score.

**Figure 3:**
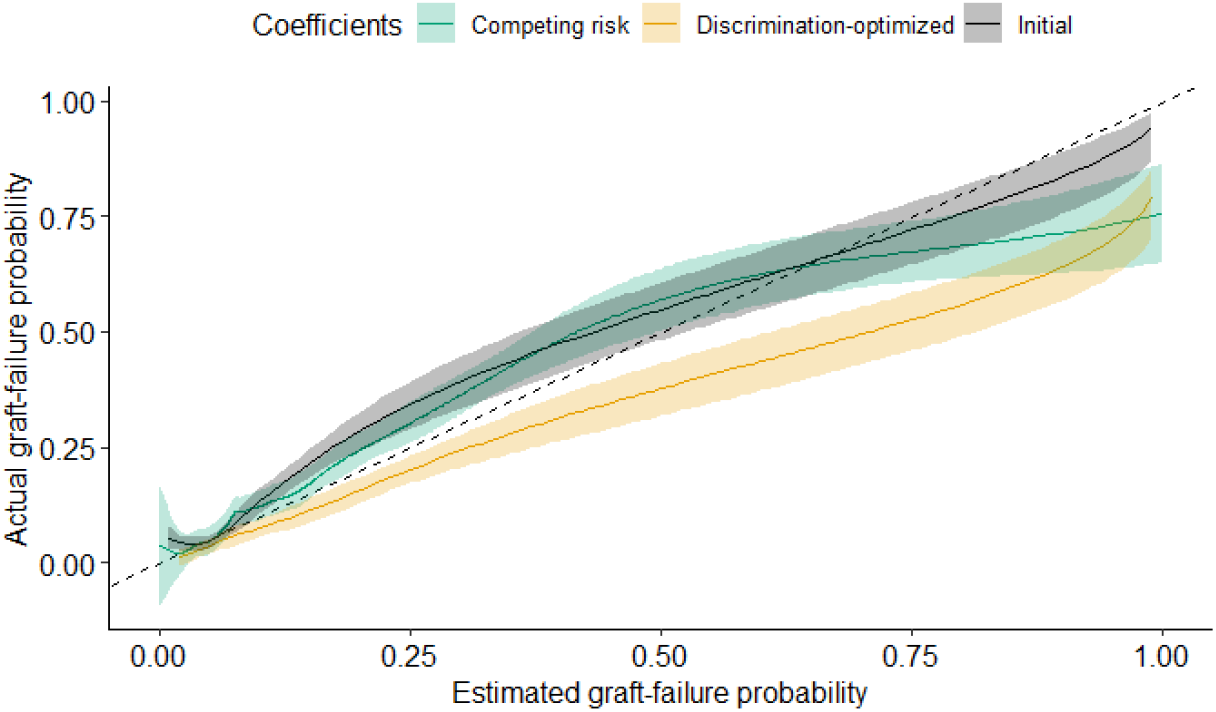
Flexible calibration curve for each possible Kidney Transplant Failure Score. Filled areas show the 95% confidence interval.

**Figure 4:**
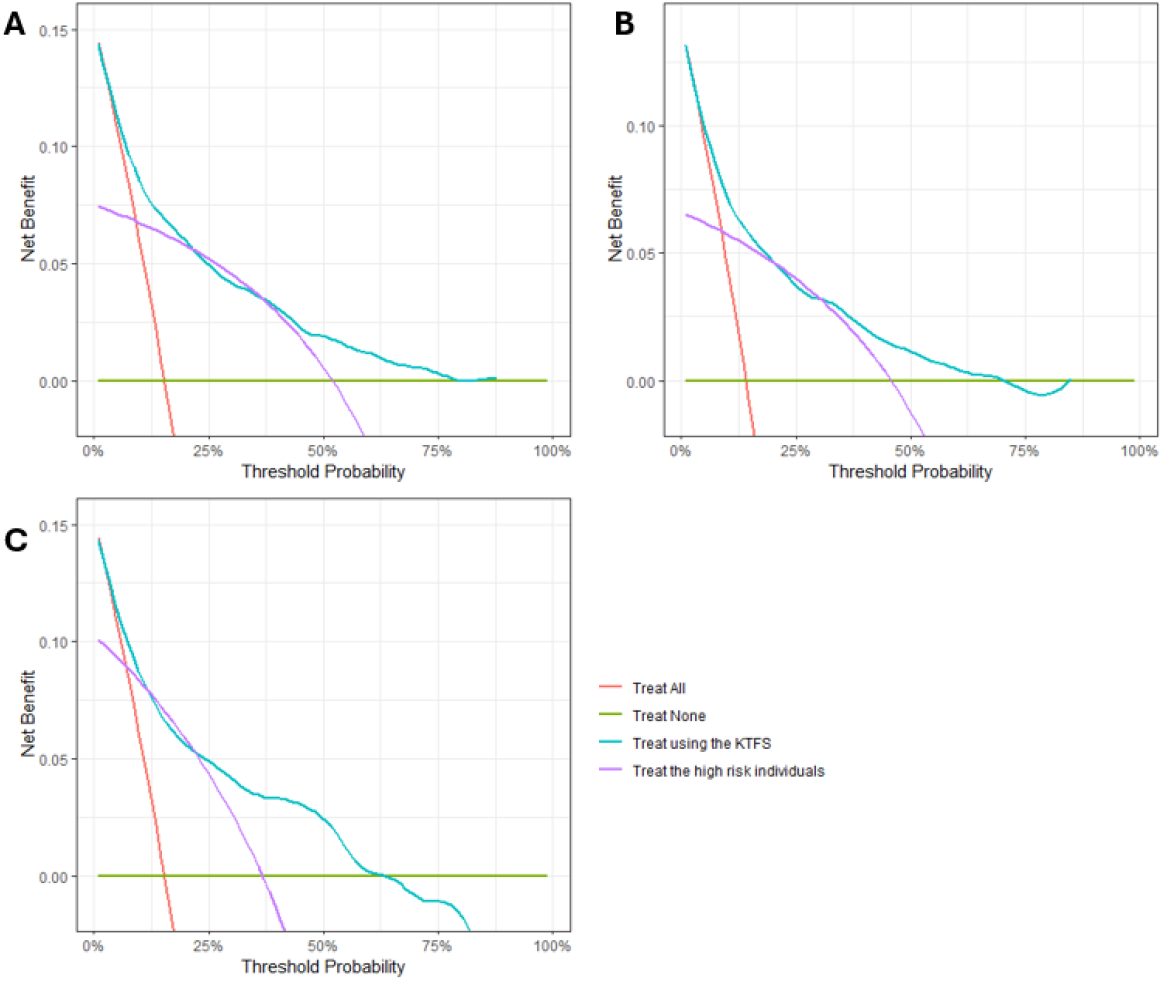
Decision curve analysis according to the possible Kidney Transplant Failure Score (KTFS) coefficients. A: Initial coefficients; B: Competing risk; C: Discrimination-optimized coefficients. The high-risk threshold was determined as a graft failure probability greater than 21.7% by the KTFS’s authors.

Country-level performances are presented in Supplementary Tables S2-4. As expected, the performance for France was similar to those of the pooled validation sample since it represents 74% of this sample. The only exception is the tAUROC of the competing risk method, which was lower for France (0.71, 95%CI from 0.66 to 0.76) than in the pooled sample (0.80, 95%CI from 0.77 to 0.84). Unfortunately, the other countries’ sample sizes are far away from the minimal sample size, yielding wide confidence intervals that preclude any firm conclusions in these populations.

## Discussion

The present study both geographically and temporally evaluated the performance of the KTFS. Our main results are that the discrimination-optimized coefficients, currently used in practice, yield a poor calibration without benefit in terms of discrimination. In contrast, the initial coefficients (i.e., not optimized for discrimination) are subject to much lower calibration drift. Notably, mean calibration did not decrease in our validation sample for the latter coefficients. Although Foucher *et al*. did not consider death as a competing event when developing the KTFS, our results show that the performance is similar when accounting for them. Our results suggest that it would be beneficial to use the initial KTFS coefficients rather than the discrimination-optimized ones currently used in clinical practice.

After performance evaluation comes impact assessment and implementation.^30^ KTFS has been used in several French hospitals since its development, and our results confirm its good performance in this population. The main obstacles to CPM clinical implementation are non-actionability (no intervention linked to the prediction), lack of safety (calibration drift not considered and hacking concerns for machine learning CPMs), and unknown utility.^31^ The impact of a KTFS-based decision (i.e., closer follow-up) is under evaluation in a randomized trial,^8^ but a cost-effectiveness analysis could complement it. However, the high-risk vs low-risk split is based on a graft failure prediction of 21.7%, selected to maximize the sensitivity and specificity.^7^ We recommend not to use this threshold because the net benefit is lower; a threshold should reflect the clinical context (and planned intervention) and may not be transportable in time and space, and continuous predictions allow for better decision-making at the individual level than risk group stratification.^32^ In addition to a closer follow-up, KTFS provides early information to anticipate a potential return to dialysis to the patient. A low probability of remaining graft failure-free during the next seven years may reinforce therapeutic adherence and involvement in the care pathway, while a high probability may reduce anxieties about an uncertain future.^33^

Some limitations must be acknowledged. First, our validation sample is mainly based on French patients (74.0%), and sample sizes from other countries preclude any firm conclusion about the generalisability. The Norwegian center performs all kidney transplantations in Norway, but additional data may be available from the other countries. Broadening the validation period is possible, but we have chosen to restrict our analysis to patients transplanted between 2010 and 2015 to leave the possibility of an 8-year follow-up while remaining as far as possible from the development period. Furthermore, we avoided transplantations during the COVID-19’s period, for which the mortality increased and the transplantation process was challenged. In France for instance, there was a kidney transplantation moratorium of 2.5 months during which kidney transplantation was not allowed, resulting in an estimated four additional months on the waiting list.^34^ For the whole year 2020, the number of kidney transplantations was reduced from 2.3% in Norway to 34.3% in France compared to 2019, resulting in more than 7500 patient life-years lost.^35^ Second, measurement approaches possibly differed between the centers and even from the development data, which may negatively impact calibration.^36^ Third, we did not include intervention-specific harms in the decision curve analysis that can reduce the net benefit. Alternative interventions could also be considered. Fourth, we replicated the original authors’ choices to only consider patients without missing predictors.^7^ Future investigations are needed to study the applicability of the KTFS in patients when at least one predictor value is missing. Fifth, no patient partner was involved in the planning or conduct of the study. Finally, we emphasize that this external validation does not mean the KTFS is “validated” in the studied populations *ad aeternam*.^37^ Other validation studies should be conducted in the future to assess temporal performance drift and use recalibration approaches if needed.^38,39^ We provided the analysis R code with both pooled and country-specific baseline hazards to conduct such recalibration in the future. Alternatively, one can consider a dynamic CPM continuously refitted over time.^40^

In conclusion, the KTFS using the initial Cox model coefficients estimated with the development sample had good performance in predicting graft failure eight years post-transplantation in this multi-country validation sample. The performance was similar when using a competing risk model. However, we warn against using the currently employed version of the KTFS – with coefficients that optimized discrimination in the original development sample – which had poorer calibration in the validation sample.

## Data Statement

All R codes are freely accessible on AC’s GitHub: https://github.com/ArthurChatton/ExtValKTFS. The EKiTE network restricts access to clinical data, as these are confidential and are subject to the General Data Protection Regulation. See Lorent et al.^12^ for information on requesting access to the data from each EKiTE center’s scientific and ethics committee. Data from the CHUM are available upon local ethics approval from HC. The protocol submitted to ethics committees was not published nor registered. However, a protocol request can be sent to AC for meta-research purposes (note that the protocol was written in French).

## Acknowledgments

We thank the people involved in the EKiTE network and the CHUM data collection process. We also thank Pr Yohann Foucher (Poitiers University, France) for sharing the original data needed to compute the Royston D-index involved in the sample size calculation and fit the cause-specific Cox model. AC was supported by an IVADO postdoctoral fellowship #2022-7820036733. HC is a Fonds de recherche du Québec senior scholar. RWP holds the Albert Boehringer I Chair. MES holds a tier 2 Canada Research Chair in Causal Inference and Machine Learning.

## Author Contributions

AC designed the study. AC, KAF, EP and HC were involved in data collection and curation. KAF, EP, and AC performed the analyses with input from MES and RWP. AC wrote the manuscript, which was revised by all coauthors.

## Supplementary materials

### Supplementary Material A: Sample size calculation

Jinks, Royston and Parmar (2015) recommended the use of either their B1 or D1 formula when (i) a measure of the Royston and Sauerbrei (2004) D-index and its standard error is available from a previous study, and (ii) a range of sample sizes is not needed. Reusing Foucher et al. (2010) data, the D-index was estimated at 2.17 (± 0.15). B1 is a significance-based sample size calculation, while D1 is a confidence-based one. Owing to the half width of a D-index difference’s confidence interval being more challenging to assume than the power, we have chosen the B1 formula:

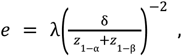

where *e* is the number of events, λ a model- and disease-specific structural constant estimated from the previous study (here 5.69), δ the posited difference in D-indexes between development and external validation (0.35), and *z* the quantiles of the standard normal laws for a posited power (80%) and significance level (5%). We assumed a one-sided test here since the D-index should theoretically decrease outside the model development sample. Alternatively, δ can be viewed as a non-inferiority margin when the test was one-sided.

Thus:

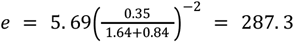

The minimum number of events is thus 288.

## Supplementary Material B: The three KTFS implementations and their coefficients

Foucher et al. fitted a Cox model to develop the KTFS, estimating the coefficients presented in the second column of Supplementary Table S1. Then, they used an optimization process to obtain the coefficients maximizing the area under the time-dependent receiver operating curve at seven years (third column). We reused their data to fit a cause-specific Cox model with death as a competing event rather than censoring its (fourth and fifth columns). Note that the graft failure’s coefficients are identical to the usual Cox model (i.e., initial KTFS), but the second model plays a role in the cumulative incidence estimation and must be estimated.

**Supplementary Table S1:** Coefficients of the different Kidney Transplant Failure Score.

|  |  |  |  |  |
| --- | --- | --- | --- | --- |
|  | Initial | Discrimination-optimized | Competing-risk<sup>a</sup> |  |
|  |  |  | Graft failure | Death<sup>b</sup> |
| Cr<sub>D</sub> | -0.76811 | -0.75072 | -0.76811 | 0.23573 |
| Age<sub>R</sub> | -0.99039 | -1.02316 | -0.99039 | 0.77335 |
| Rank | 1.07866 | 1.17295 | 1.07866 | 0.02180 |
| AR | 0.25468 | 0.22288 | 0.25468 | -0.57022 |
| Cr<sub>3</sub> | -0.00384 | 0.00188 | -0.00384 | 0.00263 |
| Cr<sub>12</sub><sup>0.5</sup> | 0.44031 | 0.41551 | 0.44031 | 0.08204 |
| Male<sub>R</sub> | -0.86668 | -0.88001 | -0.86668 | -0.08484 |
| Pr<sub>12</sub> | 0.55057 | 0.61121 | 0.55057 | 1.2690 |
| Pr<sub>12</sub><sup>2</sup> | -0.02110 | 0.04077 | -0.02110 | -0.46084 |
| Male<sub>R</sub> * Pr<sub>12</sub> | 0.50685 | 0.48605 | 0.50685 | 0.29091 |
| Male<sub>R</sub> * Pr<sub>12</sub><sup>2</sup> | -0.07623 | -0.06115 | -0.07623 | 0.02148 |
| Predictors: Cr<sub>D</sub> is 1 if donor serum creatinine is >190μmol/L (0 otherwise), Age<sub>R</sub> is 1 if recipient age > 25y (0 otherwise), Rank is 1 if transplantation rank is >2 (0 otherwise), AR is 1 if the recipient experienced at least one acute rejection episode in the first year (0 otherwise), Cr<sub>3</sub> is the recipient serum creatinine at 3 mo (in μmol/L), Cr<sub>12</sub> is the recipient serum creatinine at 12 mo (in μmol/L), Male<sub>R</sub> is 1 if the recipient biological sex is identified as male (0 otherwise), Pr<sub>12</sub> is the proteinuria at 12mo (in g/day).<br><sup>a</sup> The competing risk model is a cause-specific Cox model with a set of coefficients for the modelization of graft failure and another for the modelization of death.<br><sup>b</sup> The predictors of graft failure are used to model death, this part of the model is likely misspecified and should not be used to predict death occurrence. |  |  |  |  |

For a given patient, the graft failure probability at seven years is estimated as follows:

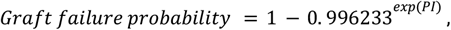

where 0.996233 is the baseline hazard for a seven years window and PI is the prognostic index calculated as follows:

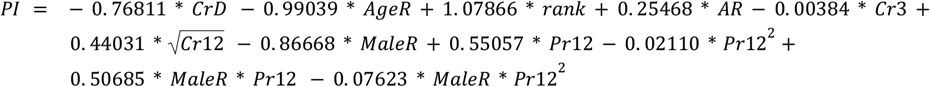

For instance, consider a male recipient named John Doe who was transplanted at 45 years old. It was his second kidney transplantation. The kidney graft cames from a donor having a last recorder serum creatinine of 85 µmol/L. In his first year post-transplantation, Mr Doe did not experience any acute rejection episode, his serum creatinine was recorded at 142 and 140 µmol/L at three and twelve months, respectively, and his twelve months proteinuria at 1.2 g/day He will have the following PI (using initial coefficients):

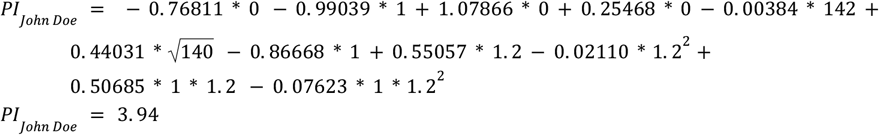

Therefore, his probability of graft failure at seven years is estimated as:

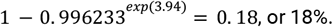

## Supplementary Materials C: Exploratory subgroup analyses at the country level

**Supplementary Table S2:** Predictive performance of the discrimination-unoptimized KTFS in the pooled validation sample and at the country-specific level.

Supplementary Materials C: Exploratory subgroup analyses at the country level.
|  |  |  |  |  |  |
| --- | --- | --- | --- | --- | --- |
|  | Pooled validation | Validation subgroups |  |  |  |
|  |  | France | Belgium | Norway | Canada |
|  | n = 3,144 | n = 2,326 | n = 255 | n = 369 | n = 194 |
| **Discrimination** |  |  |  |  |  |
| tAUROC | 0.79 (0.76-0.82) | 0.80 (0.77-0.84) | 0.74 (0.62-0.85) | 0.79 (0.63-0.92) | 0.71 (0.54-0.87) |
| **Calibration** |  |  |  |  |  |
| Mean calibration | 1.13 (1.02-1.25) | 1.16 (1.02-1.30) | 1.43 (1.01-1.90) | 1.05 (0.59-1.61) | 0.81 (0.47-1.21) |
| Weak calibration | 0.86 (0.77-0.97) | 0.83 (0.73-0.94) | 1.32 (0.88-1.77) | 1.08 (0.81-1.42) | 0.98 (0.52-1.54) |
| ICI | 0.03 (0.01-0.04) | 0.03 (0.02-0.05) | 0.05 (0.03-0.10) | 0.03 (0.01-0.08) | 0.03 (0.01-0.06) |
| **Overall performance** |  |  |  |  |  |
| sBS (%) | 20.7 (15.9-25.4) | 21.4 (15.3-27.3) | 17.0 (6.0-28.9) | 17.2 (2.5-33.5) | 13.2 (0.00-33.8) |
| Values presented: Estimated value (95% Confidence interval).<br>Abbreviations: tAUROC, area under the time-dependent receiver operating curve; ICI, integrated calibration index; sBS, scaled Brier score. |  |  |  |  |  |

**Supplementary Table S3:** Predictive performance of the discrimination-optimized KTFS in the pooled validation sample and at the country-specific level.

|  |  |  |  |  |  |
| --- | --- | --- | --- | --- | --- |
|  | Pooled validation | Validation subgroups |  |  |  |
|  |  | France | Belgium | Norway | Canada |
|  | n = 3,144 | n = 2,326 | n = 255 | n = 369 | n = 194 |
| **Discrimination** |  |  |  |  |  |
| tAUROC | 0.80 (0.77-0.83) | 0.82 (0.78-0.85) | 0.73 (0.61-0.85) | 0.76 (0.60-0.90) | 0.74 (0.57-0.89) |
| **Calibration** |  |  |  |  |  |
| Mean calibration | 0.72 (0.65-0.80) | 0.75 (0.66-0.83) | 0.83 (0.58-1.11) | 0.70 (0.40-1.08) | 0.54 (0.31-0.80) |
| Weak calibration | 0.83 (0.75-0.91) | 0.80 (0.72-0.89) | 1.06 (0.65-1.48) | 0.97 (0.64-1.34) | 0.93 (0.34-1.49) |
| ICI | 0.05 (0.03-0.07) | 0.05 (0.03-0.07) | 0.07 (0.03-0.10) | 0.04 (0.01-0.07) | 0.07 (0.03-0.10) |
| **Overall performance** |  |  |  |  |  |
| sBS (%) | 20.8 (13.4-28.0) | 22.4 (13.2-30.9) | 22.1 (4.4-37.6) | 14.7 (0.00-36.5) | 1.6 (0.00-30.0) |
| Values presented: Estimated value (95% Confidence interval).<br>Abbreviations: tAUROC, area under the time-dependent receiver operating curve; ICI, integrated calibration index; sBS, scaled Brier score. |  |  |  |  |  |

**Supplementary Table S4:** Predictive performance of the KTFS in the pooled validation sample and at the country-specific level when competing risks are taken into account.

|  |  |  |  |  |  |
| --- | --- | --- | --- | --- | --- |
|  | Pooled validation | Validation subgroups |  |  |  |
|  |  | France | Belgium | Norway | Canada |
|  | n = 3,144 | n = 2,326 | n = 255 | n = 369 | n = 194 |
| **Discrimination** |  |  |  |  |  |
| tAUROC | 0.80 (0.77-0.84) | 0.71 (0.66-0.76) | 0.77 (0.65-0.87) | 0.74 (0.59-0.86) | 0.71 (0.52-0.87) |
| **Calibration** |  |  |  |  |  |
| Mean calibration | 1.08 (0.98-1.20) | 1.12 (0.99-1.26) | 1.36 (0.96-1.80) | 0.96 (0.57-1.46) | 0.79 (0.46-1.14) |
| Weak calibration | 0.82 (0.71-0.97) | 0.80 (0.67-0.98) | 1.10 (0.66-2.86)<sup>a</sup> | 0.83 (0.51-1.30)<sup>a</sup> | 4.30 (0.43-49.2) |
| ICI | 0.03 (0.03-0.04) | 0.04 (0.03-0.06) | 0.08 (0.05-0.12) | 0.07 (0.04-0.11) | 0.07 (0.05-0.09) |
| **Overall performance** |  |  |  |  |  |
| sBS (%) | 20.1 (14.2-25.2) | 17.4 (11.4-23.7) | 13.5 (0.00-29.0) | 13.3 (0.00-31.4) | 12.3 (0.00-33.1) |
| Values presented: Estimated value (95% Confidence interval).<br>Abbreviations: tAUROC, area under the time-dependent receiver operating curve; ICI, integrated calibration index; sBS, scaled Brier score.<br><sup>a</sup>The median is presented instead of the mean due to a few extreme values from convergence issues. |  |  |  |  |  |

## Notes

### Competing Interest Statement

The authors have declared no competing interest.

### Funding Statement

AC was supported by an IVADO postdoctoral fellowship #2022-7820036733. HC is a Fonds de recherche du Quebec senior scholar. RWP holds the Albert Boehringer I Chair. MES holds a tier 2 Canada Research Chair in Causal Inference and Machine Learning.

### Author Declarations

All participants gave informed consent for research at the time of transplantation, and the University of Montreal clinical ethics committee gave ethical approval for this work.

